# Breakthrough Infections with SARS-CoV-2 in Vaccinated Students After a Festive Event – Identification of Risk Factors

**DOI:** 10.1101/2021.12.21.21266895

**Authors:** Ralph Bertram, Vanessa Bartsch, Johanna Sodmann, Luca Hennig, Engin Müjde, Jonathan Stock, Vivienne Ruedig, Philipp Sodmann, Daniel Todt, Eike Steinmann, Wolfgang Hitzl, Joerg Steinmann

## Abstract

We report an outbreak with SARS-CoV-2 breakthrough infections related to a festive event in Northern Bavaria, Germany in October 2021, with 24 of 95 participants infected. Correlation analyses among 15 interrogated variables revealed that duration at the event and conversation with the supposed index person were significant risk factors.

**Article Summary Line:** The risk of infection with SARS-CoV-2 in a vaccinated cohort associated with a private festive event was significantly increased upon conversation with the putative index person and positively correlated to the duration of stay at the event.

## Text

The severe acute respiratory syndrome coronavirus 2 (SARS-CoV-2) is the etiologic agent of the Coronavirus disease 2019 (COVID-19), classified as pandemic by the WHO on March 11, 2020 (https://www.who.int/emergencies/diseases/novel-coronavirus-2019). COVID-19 outcomes range from asymptomatic to multifactorial with fatality rates of around 1.5-2% in developed countries (https://www.statista.com/statistics/1105914/coronavirus-death-rates-worldwide/). SARS-CoV-2 is largely transmitted airborne, by aerosols and droplets (1). A viral load up to ∼3,000 SARS-CoV-2 particles has been reported in 1 L of exhaled breath of infected individuals (2), with an infectious dose as low as 10 aerosol-borne SARS-CoV-2 particles (3). Airborne transmission is positively correlated to proximity and duration of unprotected contacts to infected individuals (4). Accordingly, non-pharmaceutical prevention interventions include medical-grade face masks, handy hygiene, social distancing and ventilation of indoor spaces (5). Safe and effective vaccines have a game-changer potential against the COVID-19 pandemic. As of November 2021 four vaccines against SARS-CoV-2 have been approved by the European Medicines Agency (https://www.ema.europa.eu/en/human-regulatory/overview/public-health-threats/coronavirus-disease-covid-19/treatments-vaccines/vaccines-covid-19/covid-19-vaccines-authorised#authorised-covid-19-vaccines-section). These are viral vectors (AZD1222, AstraZeneca and JNJ-78436735, Johnson&Johnson) or lipid contained mRNA (BNT162b2, BioNTech/Pfizer and mRNA-1273, Moderna). The vaccines target the spike protein, critical for viral entry into host cells. SARS-CoV-2 variant B.1.617.2 (delta variant), which bears at least 6 characteristic mutations in spike (6), was the predominant variant in Germany in November 2021 (https://www.rki.de/DE/Content/InfAZ/N/Neuartiges_Coronavirus/DESH/Berichte-VOC-tab.html).

(Super)spreader events are serious drivers of the COVID-19 pandemic (7-10). We here report on an outbreak of SARS-CoV-2 in fully vaccinated students associated with a private festive event in October 2021 in Northern Bavaria, Germany, and shed light on factors associated with elevated risk of infection.

## The study

The event took place in Northern Bavaria, Germany, in October 20, 2021 with a duration of about 8h in a poorly ventilated room of about 85 m^2^ and a garage with less traffic. Exactly 100 students were sojourning at the location. Access was limited to those vaccinated, recovered or tested negative for SARS-CoV-2 by either PCR or antigen point-of-care test (AgPOCT) before (online Technical Appendix Table 1). Participants neither wore face masks nor practiced social distancing during the event. All further descriptions and calculations pertain to 95 persons (95%) who provided data associated with the event.

Participants were 53 males (56%) and 42 females (44%). In the aftermath of the event, 24 (25%) were diagnosed positive for SARS-CoV-2 (online Technical Appendix Table 2). All of the PCR verified isolates were of the delta type and 8 of them were sequenced. Seven were of the B.1.617.2 lineage and one of the AY.122 (B.1.617.2.122) lineage. Eight persons, all fully vaccinated, reported COVID-19 symptoms at the beginning of the event. Two of them took an AgPOCT less than 24 hours prior to the event and tested negative. Of the remaining 6, person 1.2 was diagnosed positive by AgPOCT at day 1 and PCR (cycle threshold = 24) at day 2 after the event. Thus, 1.2 as regarded as one probable index person. A proximity warning app (https://www.coronawarn.app/en/) was used by 59 of the participants and gave a posteriori alert in 52 cases (88%). Fifteen of the positively tested had used the app and 13 of them (87%) had received warning. Four of the participants had recovered from a COVID-19 infection previously. All of them had received 1 to 3 vaccinations post infection. One individual was neither recovered nor vaccinated but tested negative by AgPOCT at the day of the event. The other 90 individuals were vaccinated with 1 to 3 jabs with homologous and heterologous vaccination regimes. The periods of the last administered vaccination ranged from 15 days to 269 days (mean: 142 days, median: 141 days) before the event. Overall, 53 (56%) featured 1 or 2 risk factors for severe COVID-19. The participants attended the event for 60 to 510 min (mean 268 min, median 285 min). At least 89 guests were at the venue simultaneously with the supposed index person. Persons reported contacts with 4 – 80 other guests (mean 19.2, median 16). Figure 1 provides an overview of contact among all participants (Figure 1, panel A), or only between those tested positive after the event (Figure 1, panel B). Interactive graphs provide contact profiles for each single participant or each infected person (online Technical Appendix, URL 1 and URL 2). In all, 36 participants reported conversation with the index person. None of the infected persons required hospitalization. However, 22 reported between one and eight symptoms (online Technical Appendix Figure 1), with runny nose (n=19), headache (n=16) and loss of smell (n=14) being most prevalent. One month after the event, 10 persons reported on continued symptoms of decreased endurance or impaired sense of smell.

**Figure 1A.**
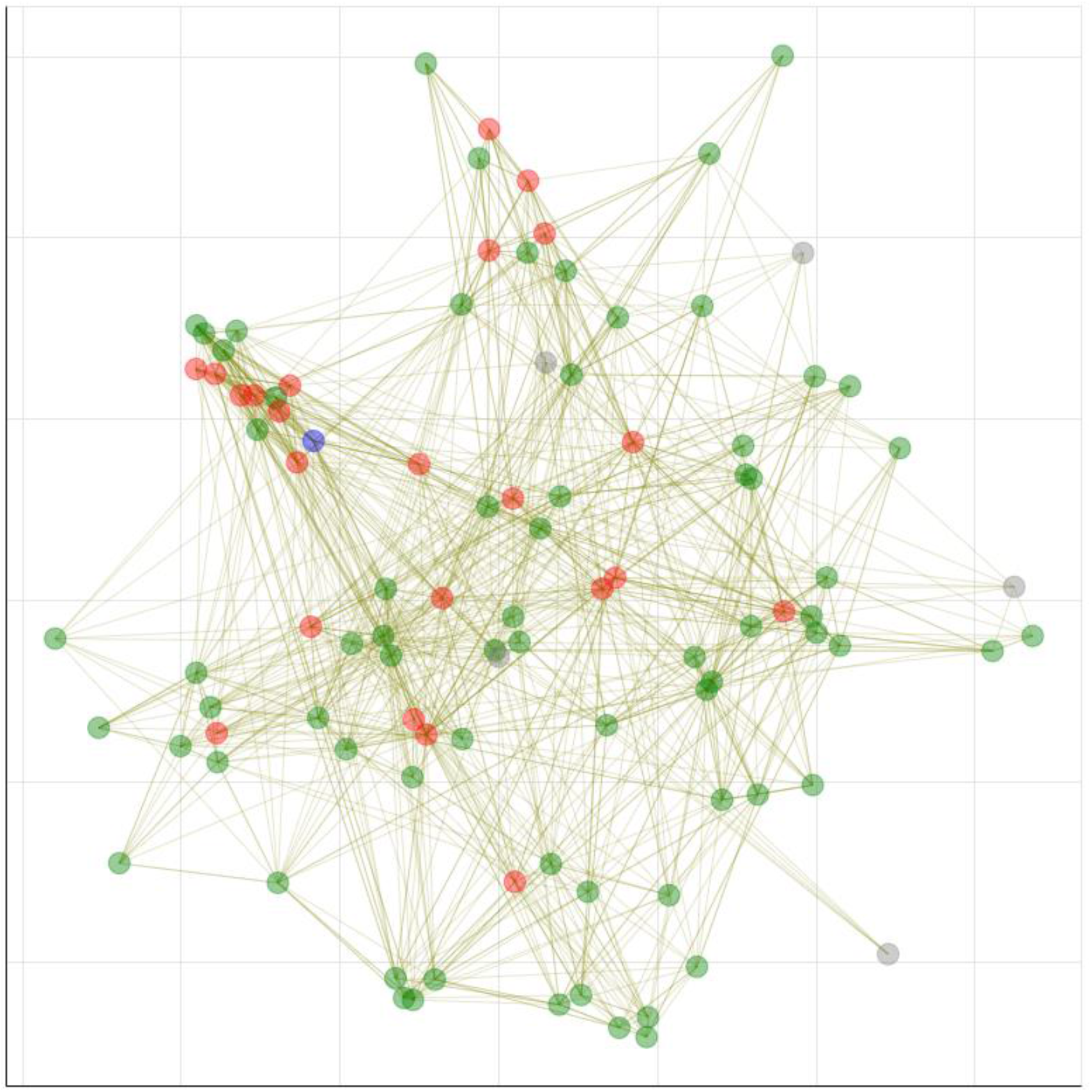
Overview of interactions between participants at the event. Each guest is represented by a node and all interactions longer than 10 minutes and a distance less than 1.5 m away are represented as edges between the nodes. The color of the nodes represents test results after the event, red indicating infection, green: no infection detected, grey: unknown. The blue node represents the putative index. To create the visualization, each node started at a random position. The spacing on the diagram and the dimensions of the axes are arbitrary.

**Figure 1B.**
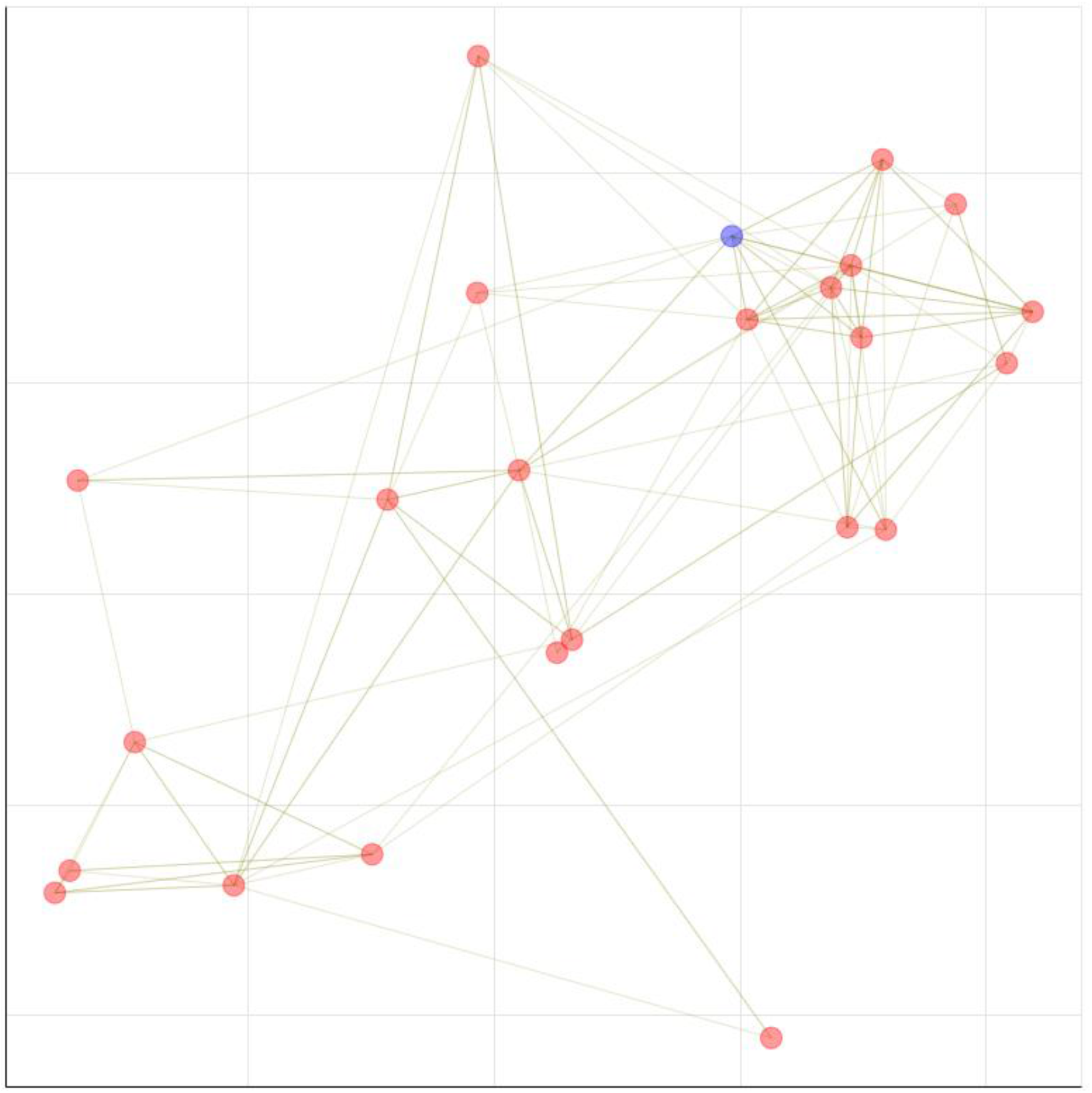
Contacts between individuals tested positive after the event. Coloring scheme as in Fig. 1A.

In search of risk factors for SARS-CoV-2 infection associated with the event, we analyzed possible correlations between interrogated variables (see supplemental material) and infection. Three correlations were significant:

i. Elongated stays at the event (Figure 2; online Technical Appendix Figure 2) (*t*-test, p = 0.0002). Notably, none of the guests that stayed 210 min or less was tested positive.
ii. Conversation with the supposed index person. Of the 24 infected persons, 13 (54%) had talked to the person. This translates to a risk of infection of 36% compared to 17% without conversation (Fisher’s Exact, two-sided, p = 0.0497). For participants staying ≥3h, we calculated risks of infection of 25% (without conversation) and 38% (with conversation with the index).
iii. The risk of infection was significantly lower upon heterologous vaccination with AZD+BNT (23%) than with AZD+AZD (60%) (Fisher’s Exact, two-sided, p = 0.0489)

In addition, the following non-significant observations were found:

- Risk of infection appeared decreased with BNT+BNT compared to AZD+AZD (Fisher’s Exact, two-sided, p = 0.14)
- Risk of infection appeared decreased with AZD+BNT compared to BNT+BNT (Fisher’s Exact, two-sided, p = 0.57)
- The duration since last vaccination appeared inversely correlated to protection (logistic regression model, p = 0.38).
- Infections of guests who shared drinking vessels were slightly increased (Fisher’s Exact test, two-sided, p = 0.24).
- The numbers of contacts was slightly increased in infected subjects (21.6 ± 12.6) vs. subjects without infection (18.4 ± 12.6; generalized log-gamma model, two-sided, p = 0.18)

**Figure 2.**
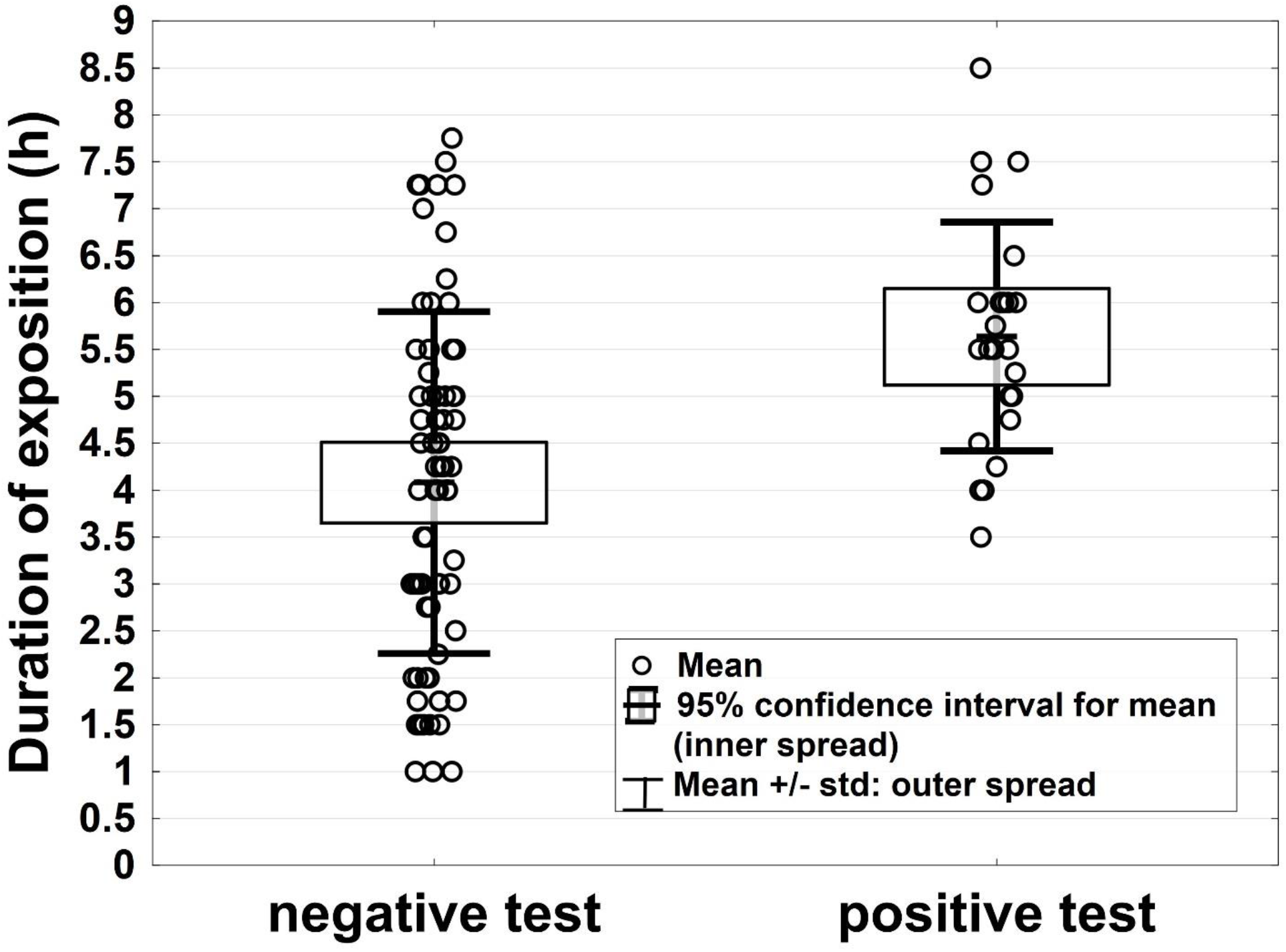
Box and whisker plot of the duration at the event in relation to a positive or negative test. Duration of exposition was significantly higher in subjects with positive tests (*t*-test, two-sided, p=0.0002)

## Conclusions

Our data show an attack rate of 25% in 96% fully vaccinated students at the event. Among the 24 infected persons, 10 declared no direct contact to the putative index person. Possible explanations for their infection are, i) more than 1 index person was at place (which may be supported by the finding of one other lineage), ii) infectious particles spread through the air within the venue and accumulated above infective threshold over time, or iii) infection did not occur at the event. Our findings regarding such infection breakthroughs are consistent with reports on waned efficiencies of the currently approved vaccines in the delta surge (11, 12). Furthermore, vaccine effects decrease with time after administration (13). A three-shot vaccination regimen with BNT162b2 as a “booster” increases efficiacy against delta (14, 15) and accordingly, only 1 of 7 individuals (14%) with three doses of vaccine became infected in our study.

Our data provide evidence that duration of stay at an event and conversation to index persons are major risk factors for COVID-19 infection during crowded indoor events with poor ventilation. Since currently available vaccines do not provide sterile immunity, high-quality testing of participants should be considered a mandatory measure to avoid SARS-CoV-2 infections at gatherings, irrespective of the vaccination status. Further studies that include larger cohorts are required to provide insights into the interplay of risk factors for SARS-CoV-2 transmission.

## Supporting information

Technical Appendix w/ Figures

interactive graph all contacts

interactive graph contacts of infected

## Data Availability

All data produced in the present study are available upon reasonable request to the authors

## Acknowledgments

We thank the Paracelsus Medical University for their support.

## Author Bio

Assoc.-Prof. Dr. Ralph Bertram is a microbiologist and a researcher and lecturer in the Institute of Clinical Hygiene, Medical Microbiology and Infectiology, Klinikum Nürnberg, Paracelsus Medical University, Nuremberg, Germany. His research interests focus on infectious diseases, drug insensitivity of nosocomial pathogens and gene regulation.

