## Supplementary material for "Breakthrough Infections with SARS-CoV-2 in Vaccinated Students After a Festive Event – Identification of Risk Factors": Technical Appendix w/ Figures

Article DOI: ____

### Appendix

### Methods

#### Description of the Outbreak Setting

The venue was a private setting, with a main room connected to a larger garage where some party activities, which required large floor space, took place. The garage was less frequented. The main venue's area was ≈85 $m^2$ (915 $ft^2$) with approximately 250 $m^3$ (8,829 $ft^3$) of airspace. To avoid noise pollution in the surrounding neighborhood, windows were closed from 10 pm onwards. Assuming an equispaced distribution of all guests in the main room, the arithmetic mean of the area per person was 0.85 $m^2$ (9.1 $ft^2$) corresponding to a personal radius of 0.52 m (1.7 ft).

#### Case Survey

A team of students from the Paracelsus Medical University in Nuremberg conducted telephone interviews with participants of the festive event that had given written consent. Data were collected in pseudonymized form, with individuals arbitrarily assigned unique numbers as identifiers. Data were compiled to an Excel file after plausibility check together with staff of the Institute of Clinical Hygiene, Medical Microbiology and Infectiology, Klinikum Nürnberg. Data collected pertained to the following variables:

- Sex (male/female)
- SARS-CoV-2-test before the event
- COVID-symptoms on the day of the event
- COVID-19 vaccination status (including date of vaccination and type of vaccine)
- Previously diagnosed SARS-CoV-2 infection

- Risk factors (see below)
- Arrival and departure at the event
- Contact to assumed index person (<1.5 m distance and >10 minutes or shorter/more distant conversation)
- Individual contacts to other guests (<1.5 m distance and >10 minutes)
- Shared use of drinking vessels
- SARS-CoV-2 infection after the event (date and kind of test)
- Symptoms up to 14 days after the event
- Symptoms 1 month after the event
- Need for hospitalization
- Use and alert of Coronavirus warning app

#### Definition of Risk Factors

We defined 12 risk factors for severe progression of COVID-19 according to the „Epidemiologischer Steckbrief zu SARS-CoV-2 und COVID-19“ https://www.rki.de/DE/Content/InfAZ/N/Neuartiges_Coronavirus/Steckbrief.html (German only), as follows: ≥70 years of age; male sex; smoker; (severe) obesity (body-mass-index>30 kg/m$^2$); cardiovascular diseases; chronic lung diseases; chronic kidney and liver diseases; psychiatric disorders; diabetes mellitus; cancer; immune suppression, trisomy 21.

#### Definition of Symptoms

We defined a case with antigen point-of-care test or PCR-confirmed SARS-CoV-2 infection as symptomatic if persons reported one or more of the following symptoms 1-8 days after the event: cough, fever (≥38.0°), common cold (blocked/runny nose), headache, sore throat, anosmia (loss of smell), or ageusia (loss of taste), shortness of breath, gastrointestinal disorders, melalgia, chest pain, tachycardia. A follow-up was conducted with those who had tested positive after the event, one month after to inquire for long-term effects.

#### Statistics and Software

Data were checked for consistency. Crosstabulation tables were analyzed using Pearson’s Chi-Square test, Fisher’s Exact test and Kruskal-Wallis test for singly ordered tables. Mann-Whitney U test and bootstrap-t tests were used to test continuously distributed variables. Logistic regression analysis was applied to test and illustrate the effect of various risk factor on the risk of infection. All reported tests were two-sided, and p-values < 0.05 were considered statistically significant. All statistical analyses in this report were performed by use of STATISTICA 13 (Hill, T. & Lewicki, P. Statistics: Methods and Applications. StatSoft, Tulsa, OK) and PASW 24 (IBM SPSS Statistics for Windows, Version 21.0., Armonk, NY).

#### Sequencing and strain assignment

Sequencing of SARS-CoV-2 isolates was performed on the Illumina platform. SARS-CoV-2 lineages were assigned using PANGO lineages (https://cov-lineages.org) (ref DOI:10.1093/ve/veab064). Characteristic mutations were furthermore identified using outbreak.info (https://outbreak.info). Multiple sequence alignments were performed using QIAGEN CLC Genomics Workbench 21.0.5 (https://digitalinsights.qiagen.com).

#### Ethics

This outbreak investigation was approved by the Institutional Review Board of the Paracelsus Medical University in Nuremberg (IRB-2021-037). Participation in the interrogation by telephone was voluntary. Written consent was obtained by all participants. Participants were informed that data protection measures were taken according to the General Data Protection Regulation of the European Union.

#### Appendix URL 1:

Interactive representation of interactions between participants at the event (as interrogated and in part supported by pictures taken at the event). Each guest is represented by a node and all interactions longer than 10 minutes and a distance less than 1.5 m away are represented as edges between the nodes. The color of the nodes represents test results after the event, red indicating infection, green: no infection detected, grey: unknown. The blue node represents the putative index. To create the visualization, each node started at a random position. The spacing on the diagram and the dimensions of the axes are arbitrary.

URL 1: “all_nodes.html“

**Appendix URL 2:**

Interactive representation of interactions between individuals tested positive after the event. Coloring scheme as in URL1.

URL 2: “only_pos“

**Appendix Table 1. Vaccination status of participants**

| Number of participants | Previous SARS-CoV-2 infection | 1st vaccination* | 2nd vaccination* | 3rd vaccination* |
|---|---|---|---|---|
| 1 | yes | JNJ | - | - |
| 1 | yes | BNT | - | - |
| 1 | yes | BNT | BNT | - |
| 1 | yes | BNT | BNT | BNT |
| 1 | no | - | - | - |
| 1 | no | JNJ | - | - |
| 10 | no | AZD | AZD | - |
| 26 | no | BNT | BNT | - |
| 3 | no | MOD | MOD | - |
| 44 | no | AZD | BNT | - |
| 1 | no | AZD | MOD | - |
| 1 | no | AZD | BNT | BNT |
| 4 | no | BNT | BNT | BNT |

Table footnotes:
*AZD, AstraZeneca; BNT, BioNTech/Pfizer; JNJ, Johnson&Johnson; MOD, Moderna

**Appendix Table 2. Selected data of infected individuals**

| Individual | Vaccination status# | Duration since last vaccine dose (d) | Number of total con-tacts | Duration at the event (min) | Kind of con-tact to index person* | Day of first posi-tive test after event | AgPOCT | PCR (cycle threshold) | Number of symp-toms |
|---|---|---|---|---|---|---|---|---|---|
| 1.2 (put. index) | 2xBNT | 178 | 12 | 270 | n/a | +1 | pos | pos (24) | 7 |
| 1.6 | 2xBNT | 136 | 12 | 315 | ++ | +4 | pos | pos (29) | 3 |

| | | | | | | | | | |
|---|---|---|---|---|---|---|---|---|---|
| 1.8 | 2xBNT | 192 | 11 | 255 | ++ | +5 | neg | pos (32) | 7 |
| 1.9 | 2xBNT | 268 | 18 | 330 | ++ | +2 | pos | neg (36) | 3 |
| 1.10 | 2xBNT | 112 | 18 | 450 | ++ | +5 | - | pos (19) | 1 |
| 1.11 | 2xBNT | 154 | 33 | 360 | ++ | +6 | neg | pos (32) | 7 |
| 1.12 | 1xJNJ | 157 | 19 | 330 | ++ | +6 | neg | pos (32) | 0 |
| 1.13 | 2xMOD | 143 | 16 | 360 | ++ | +5 | neg | pos (27) | 2 |
| 2.4 | AZD+ 2xBNT | 99 | 7 | 330 | - | +4 | pos | pos (20) | 6 |
| 2.5 | AZD+BNT | 141 | 19 | 285 | + | +6 | neg | pos (30) | 8 |
| 2.10 | none | - | 4 | 360 | ++ | +6 | neg | pos (24) | 4 |
| 2.16 | AZD+BNT | 136 | 23 | 300 | ++ | +4 | pos | pos (24) | 4 |
| 2.17 | AZD+BNT | 142 | 50 | 300 | ++ | +4 | pos | pos (20) | 6 |
| 2.24 | 2xAZD | 156 | 52 | 360 | ++ | +3 | pos | pos (18) | 5 |
| 2.27 | AZD+BNT | 141 | 19 | 360 | ++ | +5 | neg | pos (unknown) | 6 |
| 3.2 | AZD+BNT | 136 | 26 | 345 | - | +5 | - | pos (17) | 7 |
| 3.11 | 2xAZD | 165 | 20 | 330 | - | +6 | pos | pos (21) | 3 |
| 3.22 | AZD+BNT | 157 | 38 | 510 | - | +7 | neg | pos (30) | 0 |
| 3.26 | AZD+BNT | 127 | 16 | 210 | - | +5 | neg | pos (25) | 5 |
| 3.27 | AZD+BNT | 141 | 11 | 240 | - | +6 | neg | pos (34) | 6 |
| 3.28 | AZD+BNT | 141 | 8 | 240 | - | +6 | pos | pos (24) | 2 |
| 3.30 | AZD+BNT | 141 | 33 | 435 | - | +5 | pos | - | 5 |
| 3.33 | AZD+BNT | 141 | 28 | 390 | - | +6 | neg | pos (unknown) | 7 |
| 3.34 | AZD+BNT | 148 | 25 | 450 | - | +6 | neg | pos (32) | 5 |

[#]AZD, AstraZeneca; BNT, BioNTech/Pfizer; JNJ, Johnson&Johnson
[*] n/a: not applicable; ++: closer than 1.5m (4.9 ft) and for longer than 10 min; +: conversation shorter and/or more distant; -: no direct contact

**Appendix Table 3. Risk of infection relative to conversation with index person**

| | No infection | Infection | Totals |
|---|---|---|---|
| *No conversation with index person* | 48 | 10 | 58 |
| *Row %* | 82.8% | 17.2% | |
| *Conversation with index person* | 23 | 13 | 36 |
| *Row %* | 63.9% | 36.1%[1] | |
| *Totals* | 71 | 23 | 94 |

[1] Risk of infection is 2.1 times higher (relative risk) upon conversation with index, Fisher's Exact test (two-sided, p = 0.0497)

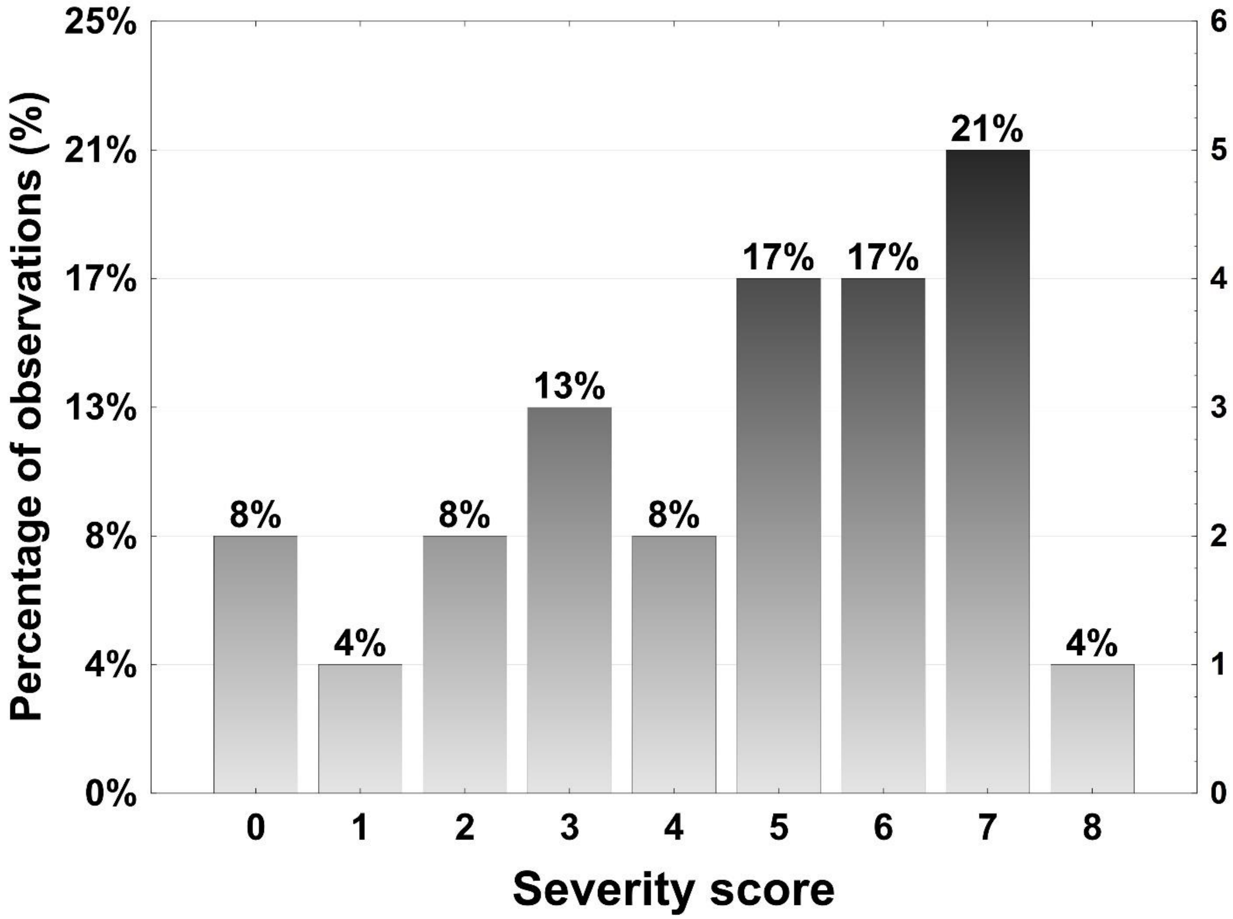


**Appendix Figure 1. Histogram of** severity of COVID-19 sickness with infected participants, according to the number of symptoms

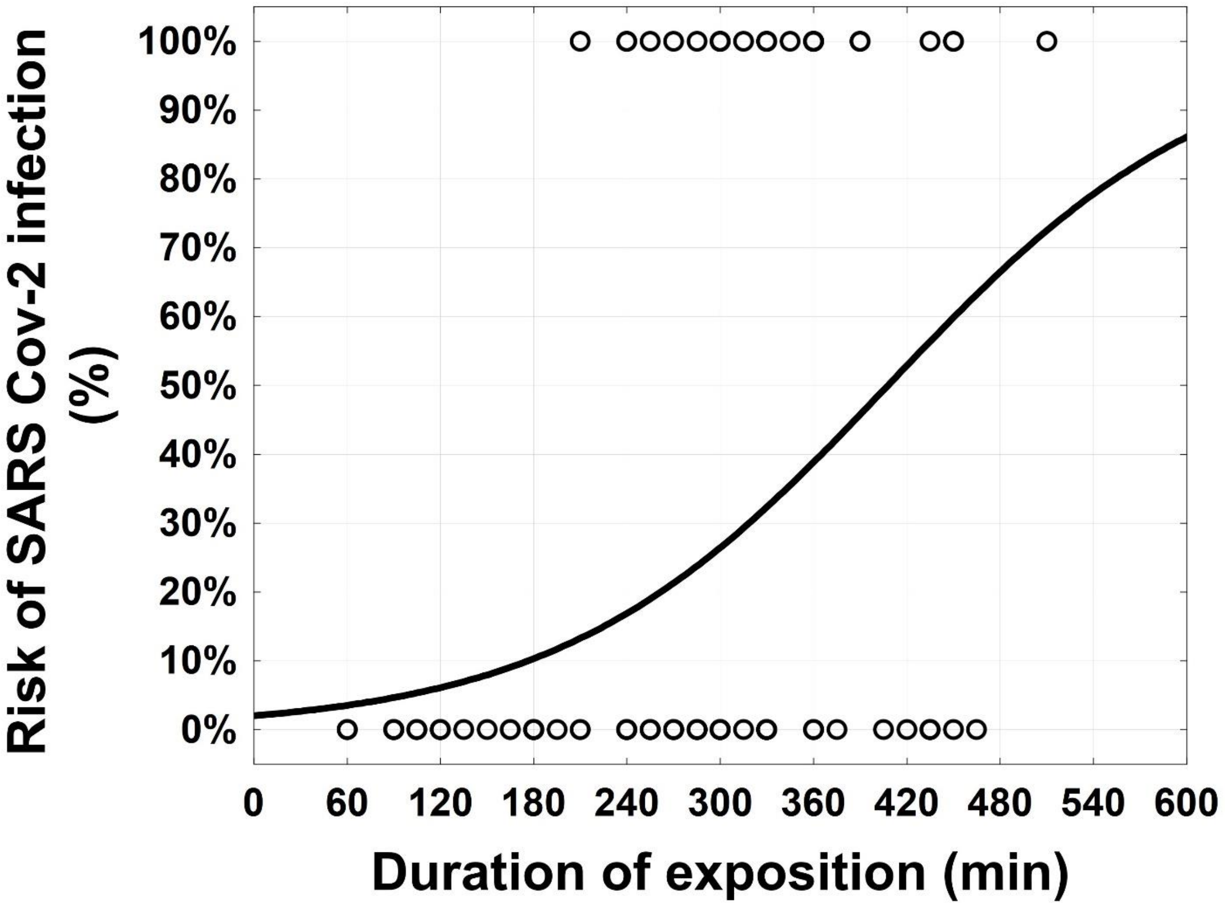


**Appendix Figure 2. Calculated risk** of infection relative to the duration of stay at the event
